# Genetic Diversity Drives the Rate and Fitness Jumps of Detectable SARS-CoV-2 Recombination

**DOI:** 10.1101/2025.10.28.25338993

**Authors:** Kyle Smith, Pranav Gangwar, Joel O. Wertheim, Yatish Turakhia

**Affiliations:** Department of Electrical and Computer Engineering, University of California San Diego, San Diego, CA 92093, USA; Department of Medicine, University of California San Diego, San Diego, CA 92093, USA

## Abstract

Viral recombination is widely considered a potent driver of adaptation, yet the epidemiological factors that influence it and the extent of its contribution to overall fitness gains of a virus remain poorly understood. In this study, we leverage three years of extensive genomic and epidemiological data collected globally for SARS-CoV-2 during the COVID-19 pandemic, combined with large-scale, unbiased recombination inference methods, to investigate recombination through an epidemiological lens. Using over 2,000 recombination events inferred from 16 million SARS-CoV-2 genomes, we show that the rate of detectable recombination is driven primarily by the standing viral genetic diversity in the population, and, to a lesser extent, by the number of infections. Our analysis highlights that >80% of detectable recombination events produce viruses with neutral or reduced fitness, indicating that recombination is consistent with the “nearly neutral theory of molecular evolution”. We also find that recombination induces larger shifts in fitness—both positive and negative—than single-nucleotide substitutions, particularly when recombinants arise from genetically divergent parental lineages. We observe that recombination between genetically divergent parents can lead to two types of epidemiological consequences. In very rare instances, <0.8% of all recombination events, it can lead to highly transmissible variants with substantial and immediate fitness gains. Additionally, seemingly neutral fitness effects arising from recombination between divergent parents, which arise more frequently (12.4% of all recombinants), may pose a subtler risk by discovering novel areas within the fitness landscape, thereby creating new pathways for rapid adaptation. These findings not only deepen our understanding of viral recombination but also have important implications for genomic surveillance efforts during a pandemic, such as to identify the high-risk periods for the emergence of recombinant variants and to guide interventions.

## I. Introduction

Recombination can create hybrid variants through the exchange of genetic material between two different genomes, providing viruses with a powerful tool for adaptation^1^. In SARS-CoV-2, recombination has been linked to the emergence of new variants with enhanced transmissibility, increased virulence, and heightened immune evasion^2–5^. Although the molecular mechanisms underlying recombination in viruses are generally well understood^6^, the extent to which detectable recombination drives the gains in viral fitness is not fully understood. Similarly, the specific epidemiological conditions that influence recombination remain poorly characterized. For example, the appearance of recombinant lineages has been shown to increase with higher levels of circulating genetic diversity^5^, which is not surprising, as genetically similar co-circulating variants are more likely to produce recombinants that closely resemble their parents, making them harder to detect. Recombination has also been observed at elevated rates during infection waves^7–9^, as higher case counts increase the likelihood of coinfections, an essential condition for recombination to occur. However, the precise relative contributions of these two epidemiological factors (i.e., circulating genetic diversity and case counts) to promote recombinants are not known, though this knowledge could help predict when new recombinant variants are likely to arise and inform intervention strategies.

In this study, we use the abundance of SARS-CoV-2 genomes to address these knowledge gaps, specifically, to study the conditions that drive the emergence of detectable recombinants and to evaluate the role of recombination in the virus’s fitness gains. SARS-CoV-2 provides an excellent dataset for this analysis, as it undergoes a modest rate of recombination^10–14^ and the majority of the COVID-19 pandemic was accompanied by reliable infection count records^15^ and an unprecedented volume of genome sequencing data^16^. Moreover, recent advancements in reliably quantifying the fitness effects of individual mutations in SARS-CoV-2^17^ and the computational ability to infer recombination at scale^10^ make this study timely and feasible. Previous large-scale studies of SARS-CoV-2 recombination^10–14^ and co-infections^7^ have not directly examined the epidemiological factors promoting recombination or the role of recombination in accelerating the pathogen’s fitness. This study focuses on *detectable* recombinants—those that introduce sufficient genetic novelty compared with their parents and are therefore better capable of altering the virus’s evolutionary trajectory.

The key findings of this study are summarized below:

1. **Genetic Diversity is the Main Driver of Detectable Recombination:** Standing genetic diversity and infection case counts both drive recombination, as sufficiently high diversity is necessary to create detectable recombinants, while high case counts help them emerge by causing more co-infections. However, analysis of 37 months of pandemic data reveals that the frequency of detectable SARS-CoV-2 recombination correlates more strongly with standing genetic diversity (Pearson R=0.78) than with raw infection counts (Pearson R=0.31).
2. **Most Recombinants are not Immediately Fitter than their Parents:** The majority (80.2%) of detectable recombination events yield viruses with neutral to negative impacts on fitness, consistent with the “nearly neutral theory of molecular evolution”. Fewer than 20% exhibit heterosis, i.e., when recombinant is fitter than both parents, and less than 9.6% reach the top 1-percentile fitness tier among circulating variants that drives global viral evolution.
3. **Recombination can Create Rare but Powerful Fitness Jumps:** Compared to single-nucleotide substitutions, recombination can generate much larger changes in viral fitness (standard deviation of 0.138 vs. 0.027). Larger genetic divergence of parental lineages enables larger jumps, both positive and negative. Though very rare (<0.8% cases), recombination between highly divergent parents can create consequential, positive fitness leaps that directly accelerate the evolution of the virus and pose heightened public health risks. An example includes the recombinant XAY’s nearly 70% fitness increase over its fittest parent.
4. **Seemingly Neutral Recombinants of Divergent Parents can Also pose Risk:** Even when the immediate fitness effect of a recombination between genetically divergent parents is nearly neutral, a more common outcome than large fitness jumps (12.4% versus 0.8% cases), recombination can place the resulting virus in a novel region of the fitness landscape, allowing it to escape “local maxima” and evolve towards even higher fitness. Such shifts can enable rapid subsequent adaptations, ultimately yielding fitness levels above both parental lineages, again underscoring why genetic diversity introduced through recombination events poses an elevated public health risk.

## II. Results

### Detectable recombination is significantly correlated with the standing genetic diversity, and, to a lesser degree, with the number of infections

We quantified the effects of standing genetic diversity and case counts on the frequency of detectable recombinant lineages using a mutation-annotated tree (MAT) comprising nearly 16 million SARS-CoV-2 sequences (Methods). Genetic diversity was quantified using the phylogenetic entropy index^18^, RIVET^19^ was used to infer recombinants, and the global SARS-CoV-2 infection counts were obtained from the JHU CSSE COVID-19 Dataset^15^ (Methods). Since the JHU CSSE COVID-19 dataset ceased to be maintained after March 2023, we restricted all our analysis to February 2023.

We explored the trends in global infection counts, standing genetic diversity, and the number of detectable recombinants inferred to have emerged each month in the 37-month period of the pandemic, from February 2020 to February 2023 (Fig. 1). To facilitate presentation, we divided the timeline into four intervals, labeled 1 through 4, highlighting distinct patterns during these time intervals (Fig. 1b).

**Figure 1:**
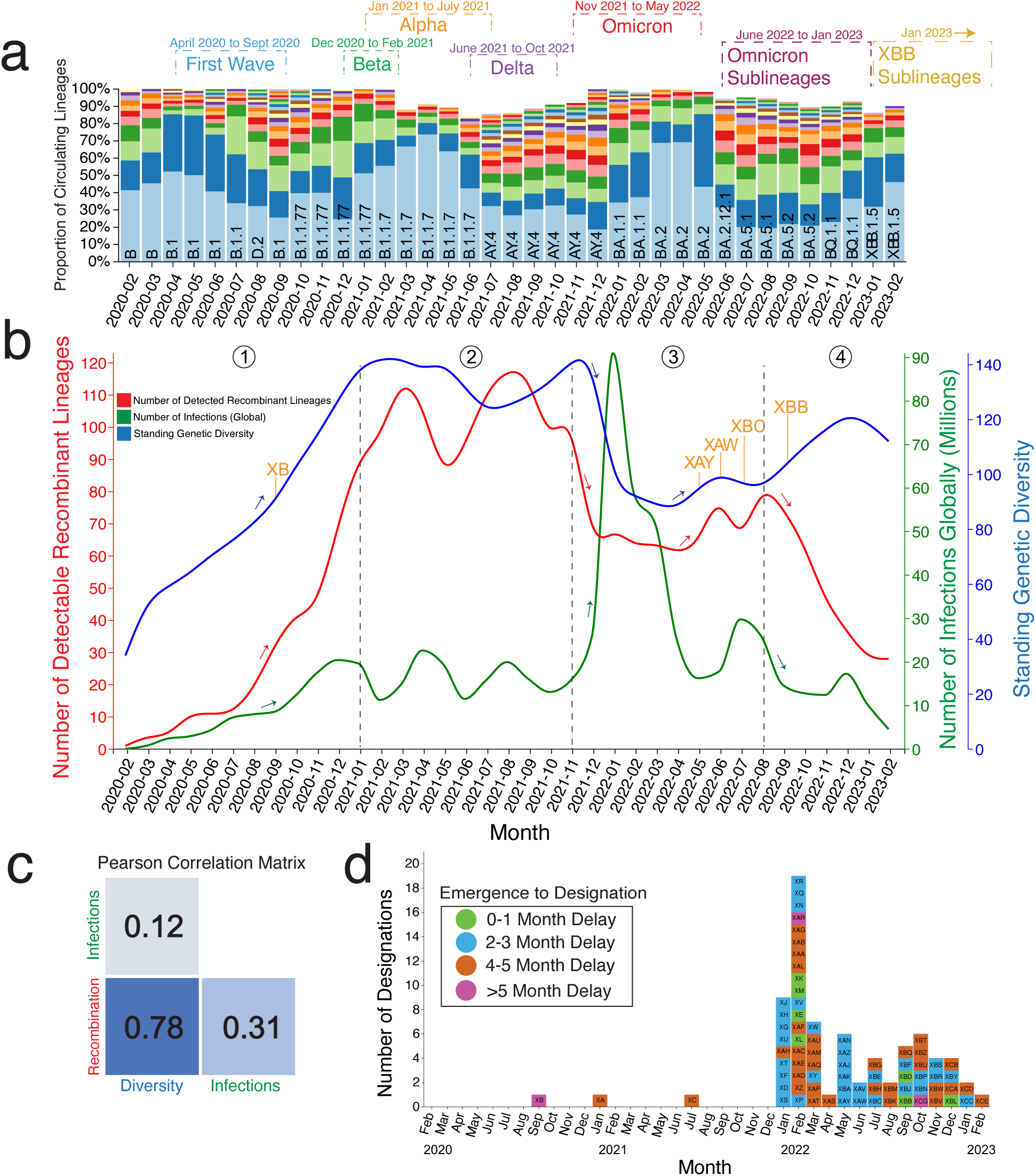
Detectability of recombinants is primarily a function of standing genetic diversity, and to a lesser extent, the number of infections. **(a)** Monthly proportion of circulating Pangolin-designated lineages, with the most predominant lineages at the bottom. **(b)** The number of detectable recombinant lineages (red), the standing genetic diversity (blue), and the number of recorded infections (green) each month during the pandemic, segmented into four intervals labeled 1-4. **(c)** The Pearson correlation coefficient matrix, which shows how the number of detectable recombinant lineages correlates with the standing genetic diversity and the number of infections. **(d)** The set of Pangolin-designated recombinant lineages plotted each month according to their respective inferred date of emergence and colored by the respective relative delay (in months) till official lineage designation.

The first interval, from February to December 2020, corresponds to the initial wave of the pandemic. This period was marked by a global rise in infection counts and the emergence of several new variants, leading to an increase in standing genetic diversity and culminating in the global dominance of the B.1.1.77 variant (Fig. 1a). The number of detectable recombinants also rose sharply during this time, peaking at nearly 70 per month by the end of the interval, even though only a single recombinant Pango lineage (XB) was identified to have emerged during this period (Fig. 1b,d). This undercount is not surprising, given that the Pango system is manually curated and prioritizes recombinants with epidemiological significance^20,21^, which was not the case for most detectable recombinants during this period. Additionally, even XB was designated months after its inferred emergence (Fig. 1d), alongside later recombinants XA and XC, as rapid tools for identifying recombinants had not been developed till then.

The second interval, from around January to October 2021, witnessed the emergence and global dominance of the named variants (e.g., Alpha and Delta) (Fig. 1a,b). During this period, global infection counts, standing genetic diversity, and the number of detectable recombinants remained relatively stable. Standing genetic diversity and the number of detectable recombinants reached their peak during this time interval (Fig. 1b); however, the ability to identify SARS-CoV-2 recombinants at scale was still limited, which may explain why only two recombinants, XB and XC, were officially designated during this period, and even these were labeled after substantial delays (Fig. 1d).

The third interval, from around November 2021 to July 2022, saw the emergence of the Omicron variant (B.1.1.529) and its many sublineages (BA.1, BA.2, BA.5, etc.) (Fig. 1a). These variants were highly infectious, resulting in a large wave with nearly a five-fold increase in global infection counts. This wave led to the extinction of most competing lineages and a sharp drop in standing genetic diversity (Fig. 1b). Notably, the number of detectable recombinants also declined during this period, suggesting that recombination detectability is more strongly correlated with genetic diversity than incidence (Fig. 1c). By the end of the interval, case counts had returned to similar levels as the previous interval, while standing genetic diversity and the number of detectable recombinants were stable. Heightened interest and improved ability to identify SARS-CoV-2 recombinants, particularly between Omicron and Delta lineages, resulted in several designations, such as XAY, XAW, and XBC, during this period with shorter delays (Fig. 1d).

The final interval, from August 2022 to February 2023, was characterized by the continued dominance of Omicron sublineages, several of which emerged during this period, causing a slight increase in standing genetic diversity. Notably, the XBB lineage emerged, was rapidly designated (Fig. 1d), and became the first recombinant whose sublineages, like XBB.1.5, achieved global dominance (by abundance) in infections (Fig. 1a). However, as case counts declined to levels comparable to the first wave (pre-2022), the number of detectable recombinants also dropped sharply, indicating that the number of recombination events is also associated with infection counts (Fig. 1c).

Across the four intervals, we found a significant positive correlation observed between the number of detectable recombinants and standing genetic diversity (Pearson R = 0.78) and a more moderate correlation with infection counts (Pearson R = 0.31) (Fig. 1c). Our findings suggest that genetic diversity is the primary driver of detectable recombinant emergence. However, in time periods where genetic diversity is already high, infection counts may become the limiting factor for recombinant emergence. We note that this pattern is not readily apparent through Pango-designated recombinants (Fig. 1d, see Methods) and required a comprehensive, unbiased approach to recombinant inference, such as with RIVET, to uncover.

### Detectable recombinants are rarely within the top 1% of circulating fitness

To assess the role of recombination in accelerating the fitness increase of SARS-CoV-2, we used PyR_017_ to compare the fitness of all detectable recombinants emerging each month with the average circulating fitness: the mean fitness of all globally circulating sequences during that month (Fig. 2a, Methods). We compared these values to the top 1-percentile fitness of the circulating sequences (highlighted region in pink, Fig. 2a), which yielded insightful observations. As expected^17,22^, the average circulating fitness of SARS-CoV-2 increased over time (Fig. 2). Notably, we find that it typically takes only 1 to 2 months for the top 1% fitness value in a given month to become the average fitness of all globally circulating sequences (Fig. 2a). This observation affirms that the fittest sequences swiftly move on to become the predominant circulating lineage and are the ones contributing to the overall fitness increase of the virus over time.

**Figure 2:**
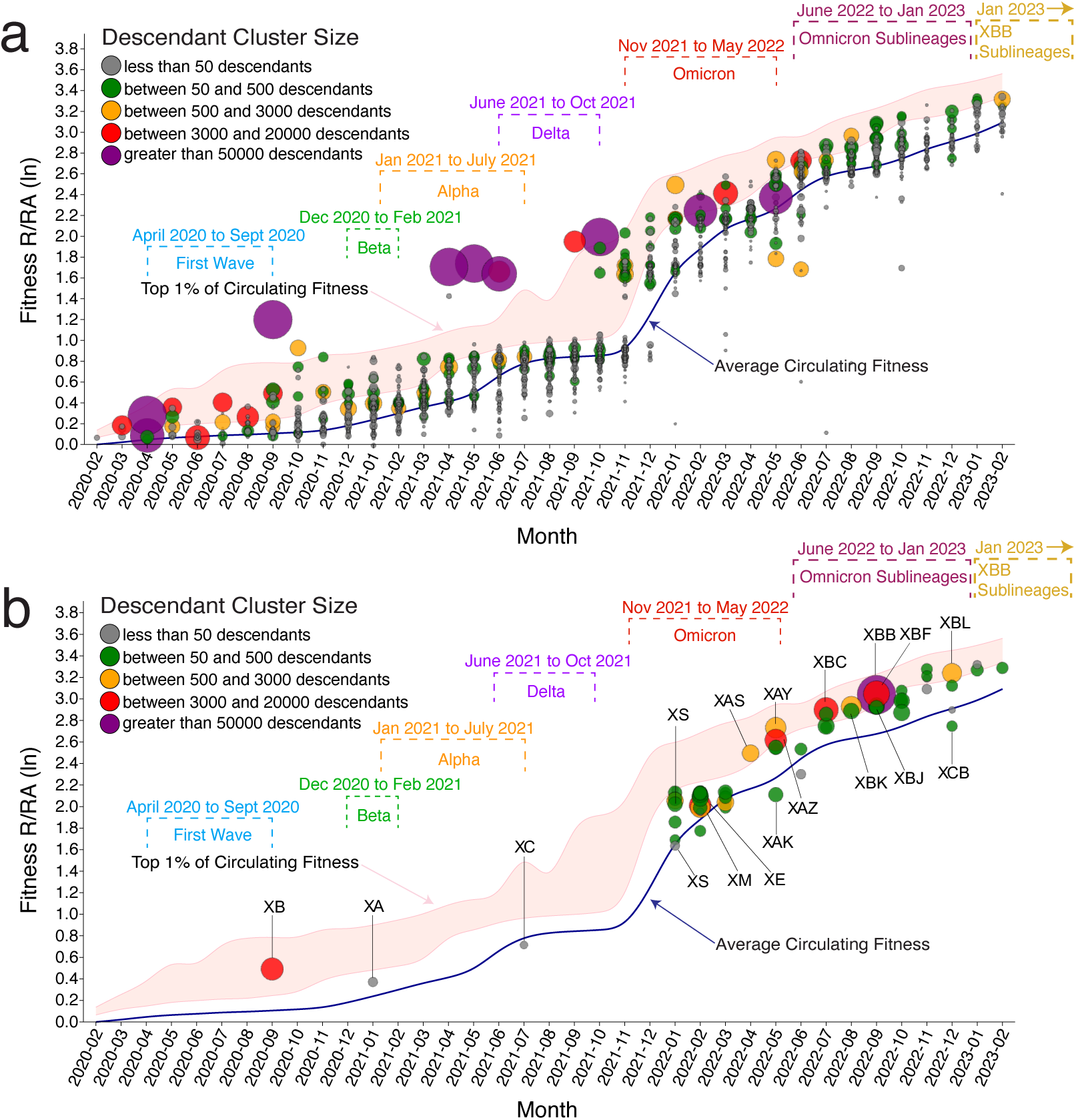
Fitness of RIVET-inferred and Pango-designated recombinants over the course of the pandemic. (**a**) The relative fitness (in natural logarithm scale) of RIVET-inferred recombinants shown according to their respective inferred emergence dates between February 2020 and February 2023. Each recombinant lineage is colored and scaled according to the size of its descendant cluster in the UShER MAT. The blue curve denotes the average circulating fitness of all samples sequenced during the respective month, and the pink region shows the top 1-percentile fitness of all samples for that month. Circles outside the pink shaded region represent recombinant nodes for which the Chronumental-inferred dates (Methods, Fig. S1) were substantially earlier than those of their descendant samples. (**b**) The corresponding plot for the set of Pango-designated recombinant lineages over the same time interval.

We observe that fitter RIVET-inferred recombinants tend to form larger descendant clusters (reflected in the size of the corresponding circles in Fig. 2a). We also observe that most recombinants have fitness values concentrated near the average fitness circulating in the corresponding month, with less than 9.6% achieving fitness within the top 1% of circulating sequences. These observations suggest that, compared to the evolutionary trajectory of the virus through vertical descent, the immediate fitness jumps achieved directly through recombination events are likely not an outsized contributor to the fitness increases in the virus. However, this finding does not rule out the possibility that recombination indirectly influences fitness gains over time—an aspect that we explore in a subsequent section.

Once again, the observations that the concentration of recombinant fitness near the average and rarely within the top 1% of circulating fitness are less evident when only Pango-designated recombinants are considered (Fig. 2b), as the Pango system prioritizes naming recombinants that have the greatest epidemiological impact. Consequently, a larger fraction (20%) of Pango-designated recombinants fall within the top 1% of circulating fitness.

### Most recombination events have a neutral to slightly deleterious effect on viral fitness

We then examined how recombination events influence fitness relative to the recombining parents. For most detectable recombinants (64.9%), the PyR_0_ fitness values are between those of its two parents, with fewer cases exhibiting fitness lower (15.4%) or higher (19.8%) than both parents (Fig. 3d, Methods). We chose to focus on the comparison to the fitter parent, because it highlights whether recombination produces offspring that are fitter than the circulating parental sequences, a phenomenon sometimes referred to as heterosis^23^. Analyzing the distribution of recombinant fitness normalized to the fitter parent’s fitness (Fig. 3b), we observe that recombination, on average, has a slightly deleterious effect, with recombinant fitness averaging 93.7% of the fitter parent’s fitness. However, most recombinant virus fitness values cluster around 1, indicating near-neutral effects of recombination (Fig. 3b,c,d). Thus, a recombination event, like a point mutation, typically has a neutral to slightly deleterious impact on viral fitness.

**Figure 3:**
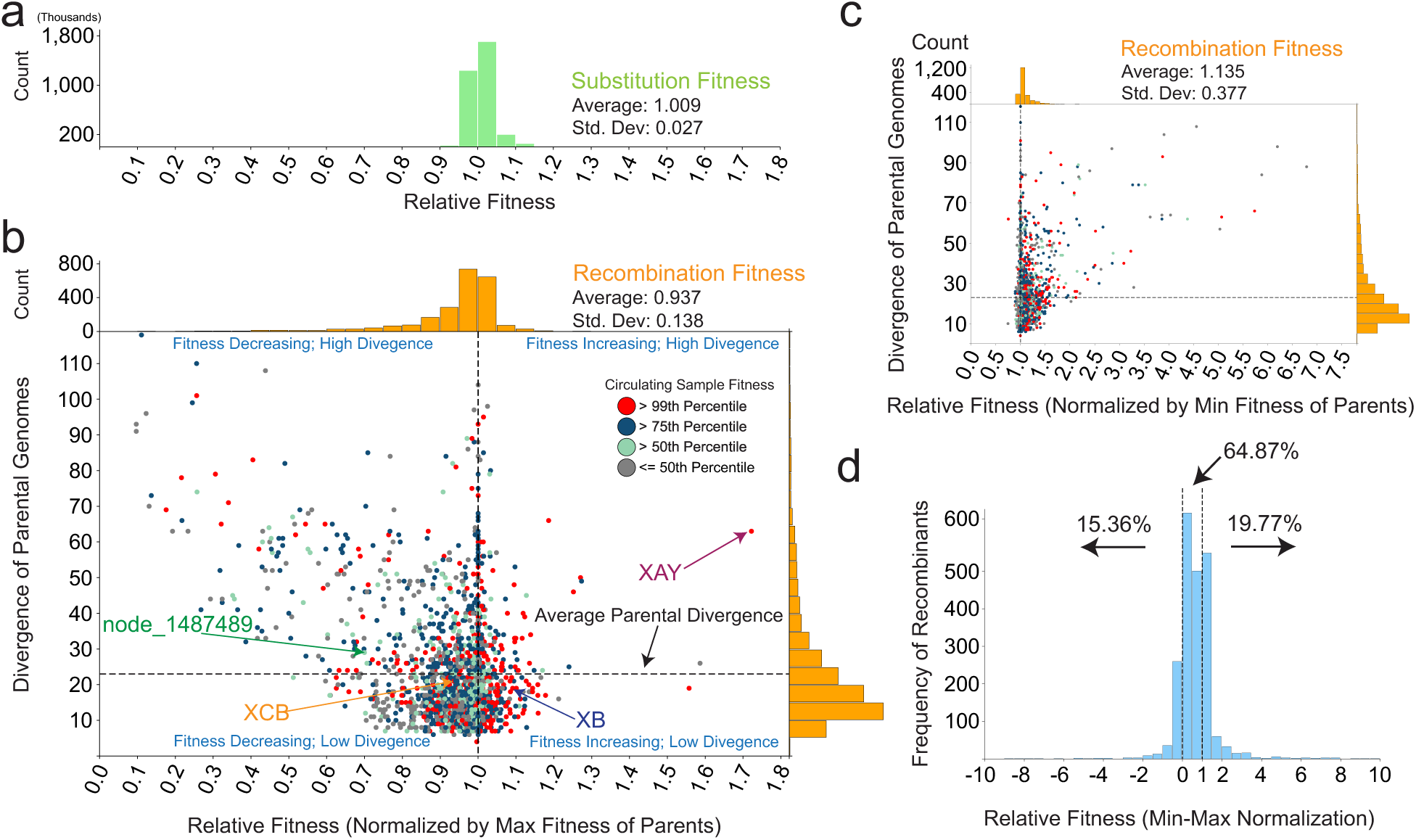
The relationship between recombinant fitness and divergence of parental genomes. **(a)** Fitness distribution of single-nucleotide substitutions acquired through the typical vertical mode of evolution. **(b)** The relationship between the recombinant’s fitness normalized to its fitter parent versus the divergence (in number of sites with differing alleles) between the two parental genomes. Each of the four quadrants is labeled with a case-study recombinant, three of which were designated by Pangolin, and examined in more detail in Fig. 4. Each recombinant in the scatter plot is colored according to its fitness ranking among circulating samples during the month it was inferred to have emerged. (**c**) Shows the same scatter plot as panel b, except with the recombinant fitness normalized to the fitness of the less fit parent. (**d**) The distribution of recombinant fitness advantage compared to the parental sequences, with each recombinant’s fitness value rescaled using min-max normalization^26^ with respect to their parental fitness range.

Most detectable recombinants appear to originate from genetically similar parental sequences, resulting in relatively small fitness jumps, with negative fitness effects—where the recombinant is less fit than the fittest parent—being more frequent (Fig. 3b). Whereas the spread of fitness jumps expands with greater parental divergence, the majority of recombinants remain clustered around the nearly neutral fitness region (i.e., close to 1). Recombination that occurs between sequences that are already below the median circulating fitness is unlikely to produce a competitive recombinant, occurring in fewer than 6.8% cases. One such example is the XCB recombinant, between the BQ.1 and BF.31.1 lineages, where several highly ranked mutations are absent in the recombinant child sequence, including an ORF14:S4L mutation carried by the BF.31.1 parent sequence (Fig. 4a). Despite the later acquisition of a A262S spike protein mutation, which was suggested to have increased ACE2 utilization^24^ and a reduction in the efficacy of treatment with monoclonal antibodies^24,25^, this lineage remained predominantly confined to a relatively small local transmission cluster in Chile.

**Figure 4:**
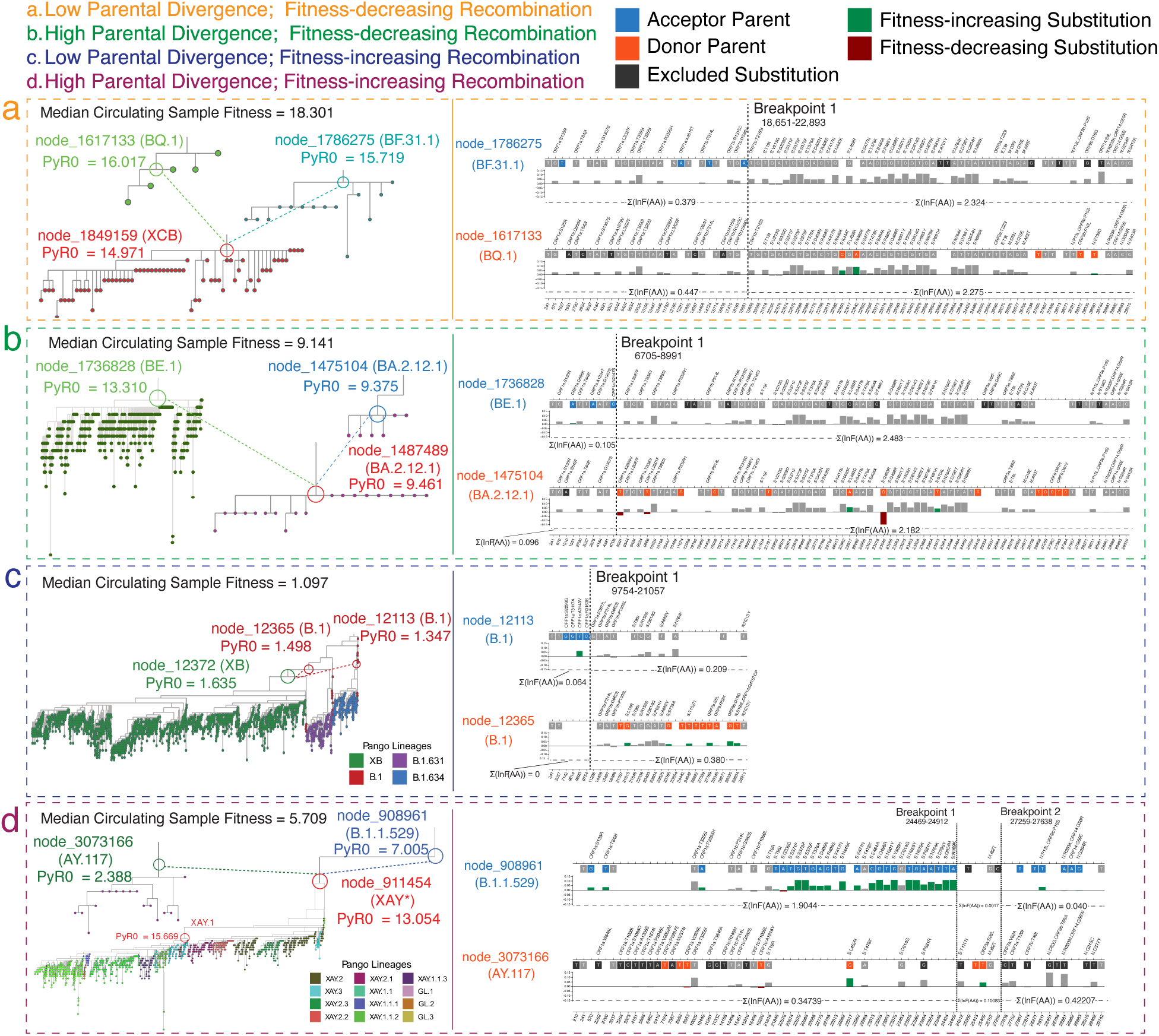
Case studies of SARS-CoV-2 recombinants. (**a**) ‘Low Divergence, Low Fitness’ case. On the left panel, the approximate phylogenetic tree topology (Methods) highlights a RIVET-inferred recombinant node equivalent to the Pangolin XCB recombinant lineage root resulting from BQ.1 (donor) and BF.31.1 (acceptor) parental lineages. The PyR0 fitness for both parental lineages and the recombinant node, as well as the median circulating fitness at the time of the recombination event, are labeled. On the right panel, the acceptor (blue) and donor (orange) protein-coding, non-synonymous single-nucleotide substitutions are shown. The vertical dashed lines denote the inferred breakpoint intervals for the detected trio. The relative fitness (PyR0) of each amino acid mutation is shown on the track below each nucleotide change, with fitness-increasing (green) and fitness-decreasing (red) mutations highlighted. (**b-d**) Similar to (a) but highlighting the ‘High Divergence, Low Fitness’ case using a RIVET-inferred recombinant node labeled node_1487489 in the UShER MAT, the ‘Low Divergence, High Fitness’ case using a RIVET-inferred recombinant node equivalent to the Pangolin XB recombinant lineage root, and the ‘High Divergence, High Fitness’ case using the closest RIVET-inferred recombinant node (one branch above) to the Pangolin XAY lineage root.

Though less common, recombinants that exhibit enhanced fitness compared to both parents tend to be among the top 1% or top 25% of the circulating sequences. We observe several instances where two genetically similar, high-fitness parental sequences recombine to produce a highly fit recombinant. One such example is the Pangolin-designated XB recombinant, which was the result of two fit B.1 sublineages (Fig. 4c), producing a sizable descendant cluster that successfully spread throughout Central and North America in late 2020.

### Recombinants hold greater potential for large jumps in fitness than substitutions

We then analyzed the fitness effects of recombination and substitutions, as inferred from UShER’s MAT, and gained two additional insights. First, we found that most substitutions cluster in the nearly neutral region (fitness close to a value of 1), with a mean fitness value of 1.009. Similar to recombination, highly deleterious substitutions are rapidly purged from the population and are thus largely absent in the MAT. Although recombination more frequently reduces fitness relative to the fitter parent, most observed substitutions slightly increase the fitness relative to its parent. However, this trend reverses when a recombinant is compared to its less fit parent (Fig. 3c).

Second, we found that compared to substitutions, recombination events can lead to greater swings in fitness, both positive and negative. This pattern is evident in the substantially higher standard deviation of 0.138 for the fitness changes through recombination events, compared with 0.027 for substitutions (Fig. 3a,b).

### Increasing divergence of parental sequences broadens the spread of recombinant fitness jumps

We expect the magnitude of fitness jumps to increase with the divergence of parental sequences, as diverse parents can generate hybrids with markedly distinct genotypic configurations. To examine this relationship, we explored the fitness of the resulting recombinant normalized to the fitness of the fitter parent as a function of genetic distance (Hamming distance of their corresponding single-nucleotide alleles) between the parents (Fig. 3b). Relative fitness values above 1 (right of the dotted vertical line) indicate that the recombinant surpassed both parents in fitness (heterosis), whereas values below 1 (left of the line) signify that the recombinant was less fit than at least one parent. As expected, the V-shaped spread of the data points on the scatter plot suggests that increasing parental sequence divergence broadens the spread of fitness jumps observed in recombinants.

To better characterize the fitness of recombinant viruses, we organized our analysis into four quadrants defined by two key factors: (1) Parental Genetic Divergence—whether the genetic divergence between the two parents is low or high, and (2) Recombinant Fitness—whether the recombinant is more or less fit than its parents. Using case studies from Fig. 4, we examine each of these four scenarios below.

#### Low Divergence, Low Fitness (52.2% cases)

The most frequent scenario involves two closely related parents that recombine to produce a recombinant with slightly lower fitness (Fig. 3b). An example is the Pango lineage XCB (Fig. 4a), which arose from a single recombination event where a single breakpoint caused the less fit segment from each parent to be joined. The resulting XCB lineage was slightly less fit than both of its parents, as well as the median circulating viral fitness at that time (Fig. 4a). As a result, XCB went extinct without spreading widely. This scenario is most common and least consequential from an epidemiological perspective.

#### High Divergence, Low Fitness (28.1% cases)

Recombinants from highly divergent parents are less common, and when they do occur, they often result in lower fitness (Fig. 3b). For example, the recombinant at MAT node labeled node_1487489 (undesignated in Pango, Fig. 4b) experienced a sharp drop in fitness compared to its more fit parent, BE.1. This drop was primarily due to acquiring the Q493R mutation in the receptor-binding domain (RBD) of the Spike protein from parental lineage BA.2.12.1, a mutation with a large negative PyR_0_, the reversion of which has been shown to have an increased ACE2-S1-RBD affinity in several highly-infectious Omicron sublineages, including BA.4 and BA.5^27^. Although this recombinant was slightly fitter than the median circulating virus, it went extinct quickly, likely because much fitter lineages (like BE.1) were co-circulating. This scenario is also not epidemiologically concerning (explaining why it is rarely featured in Pango designations), and its fitness drops can be even higher than those of the first scenario.

#### Low Divergence, High Fitness (13.6% cases)

A rare scenario involves low-divergence parents creating a recombinant with higher fitness, such as the Pango lineage XB (Fig. 4c). In this case, the recombination event combined the fitter segments from each parent. Since both parents were already relatively fit, the resulting recombinant had a significant advantage over circulating lineages and spread rapidly, leading to many descendants (Fig. 4c, left). This scenario could be concerning, but it is rare that a combination of segments from closely related viruses can produce large fitness increases.

#### High Divergence, High Fitness (6.1% cases)

The rarest scenario is when two highly divergent parents recombine to create a remarkably fitter recombinant. This combination can lead to significant fitness jumps, implying more transmissible variants that can increase the public health burden. The XAY recombinant, which emerged during the co-circulation of Delta and Omicron, serves as a prominent example (Fig. 3b, 4d). XAY acquired advantageous genomic segments from its genetically diverse parents, Omicron lineage B.1.1.529 and Delta lineage AY.117, which differed at 63 sites (Fig. 3b, 4d). This recombination resulted in an offspring with a nearly 70% fitness increase over the fitter parent, placing it in the top 1-percentile of all circulating sequences (Fig. 3b, 4d). This advantage allowed it to spread rapidly and spawn several descendants and sub-lineages (e.g., XAY.1 and XAY.1.1, Fig. 4d). A recombinant with fitness at least 10% higher than that of the fitter parent was observed only in 0.79% of detectable recombination events, indicating that although such events can directly accelerate viral evolution and pose a serious public health concern, they are exceedingly rare.

### Seemingly neutral recombination events from diverse parents may speed up evolution by enabling a shift to a new fitness regime

Although recombination between highly divergent parents tends to produce larger swings in fitness, most resulting detectable recombinants still fall within the near-neutral range (Fig. 3b). For example, approximately 12.4% of recombinants arise from highly divergent parents and have normalized (to fitter parent) fitness between 0.98 and 1.02 (i.e., within the near-neutral range). A closer look at such cases, which are often overlooked and remain undesignated in the Pango nomenclature system, reveals another way in which recombination can indirectly accelerate viral evolution. For example, the MAT node labeled node_240960 is inferred to be a recombinant of two generically distant parental sequences belonging to B.1.526 and AZ.2 lineages and differing at 42 sites (Fig. S3). The recombinant’s initial PyR_0_ fitness (1.570) is slightly lower than that of its fitter parent (1.572), yet a single branch after the recombination event, the PyR_0_ fitness at node_240980 achieves a higher fitness than both parents (1.63). Another such example is that of node_596267, which is inferred to be a recombinant of two genetically similar lineages, BA.1 and BA.1.20, differing at 14 sites (Fig. S4). Here too, the recombinant’s initial PyR_0_ fitness (5.35) is slightly lower than that of its fitter parent (5.42), yet the recombination event introduced substantial genomic novelty, particularly in the Spike protein-coding region (Fig. S4), placing it in a distant, previously unexplored region of the fitness landscape. In this new region of the fitness landscape, vertical evolution could proceed along mutational paths unavailable to either parent, potentially enabling the lineage to cross a fitness valley and reach a higher peak. Indeed, within two short descendant branches, the recombinant acquired the S:R346K mutation, the third-highest ranking mutation in the PyR_0_ model, raising its fitness to 5.82, far surpassing the fitness of both parental lineages (Fig. S4). This jump triggered rapid expansion of the recombinant lineage, resulting in 734 additional descendants (1,518 total), compared with 43 and 30 descendants for the parental lineages (Fig. S4).

These two examples highlight that immediate fitness gain is not the sole pathway through which recombination accelerates adaptation. Particularly when involving genetically divergent sequences, recombination can also generate novel genetic backgrounds that enable vertical evolution to achieve substantial fitness gains through future mutations. In the field of optimization, this process resembles strategies like random restarts, which introduce diversity to help a search algorithm escape a local optimum and find better solutions elsewhere^28^.

## III. Discussion

We performed a large-scale study investigating the epidemiological factors that promote recombination and the role of recombination in accelerating viral fitness gains. Our study reveals that genetic diversity contributes more to the emergence rate of detectable recombinants than case counts. However, a combination of high diversity and case counts, as occurred during the co-circulation of Delta and Omicron in early 2022, creates ideal conditions for epidemiologically consequential recombinants to arise.

Our study finds that the fitness effects of recombination align with the predictions under the “nearly neutral theory of molecular evolution”^29,30^, which posits that most standing genetic diversity has minimal selective effects, tends towards mostly negative fitness consequences, and can be shaped by genetic drift that depends on the population size. Although prior studies have validated this theory in the context of point mutations (substitutions)^31,32^, our findings reveal that the nearly neutral theory also generalizes to recombination. Although we observed that most recombination events have small, slightly negative effects on fitness, rare recombination events can produce substantial gains in viral fitness. We show that, compared to substitutions, recombination events can cause larger swings in fitness values, both gains and losses. Although this trend could be expected since recombination simultaneously affects multiple genomic sites, it underscores the unique epidemiological risks posed by recombination as compared to vertical mutations, since recombination can lead to more dramatic leaps in viral fitness. Lastly, we also note that the genetic diversity of circulating variants not only has the potential to contribute to large fitness swings but can also complement vertical evolution by enabling lineages to escape local fitness peaks and achieve higher eventual fitness.

This study was made possible by three key developments. *First*, COVID-19, which was the first pandemic in the post-genomic era^33^, and was marked by unprecedented genome sequencing efforts and relatively reliable infection tracking worldwide, which together provided the data necessary for this analysis. *Second*, recent years have also witnessed the emergence of scalable computational tools capable of handling pandemic-scale data, for inferring phylogeny (UShER^34^), dating nodes (Chronumental^35^), inferring recombinants (RIVET^19^), and accurately estimating the fitness effects of different variants (PyR_017_). These advances have been critical to facilitating this study, though not without limitations. For example, well-documented global disparities exist in the sequencing rates, metadata quality, and infection reporting^36^. We mitigate this bias by focusing our analysis on broad global trends, which are likely to be robust against such regional inconsistencies. Similarly, most recombination likely occurs between variants that are genetically nearly identical. These recombinants likely go undetected via RIVET, but such events are also of limited epidemiological relevance because they do not directly produce novel strains. Despite its stringent quality filters, RIVET identified recombinants more comprehensively than those designated under the Pango system (2,266 vs. 75 in the time interval considered), including several that may not have caught the attention of human experts (Fig. 2). *Third*, our analysis relied on PyR_0_, as this is the only available model for SARS-CoV-2 fitness that considers mutations across the entire genome. This approach was essential to study the effects of genome-wide recombination breakpoints. A limitation of PyR_0_ is that it does not model epistasis, because it assumes the independent contribution of each mutation to viral fitness. However, we found that PyR_0_’s fitness estimates are well-correlated with those from Spike-protein-based models that capture epistasis, like CoVFit^37^ (Fig. S2, Methods). This observation suggests that PyR_0_ accurately captures the first-order fitness effects. Although higher-order effects may exist, they do not seem large enough to alter the broad conclusions drawn in this study.

In summary, this study provides an in-depth analysis of the forces shaping viral recombination. By clarifying the interplay between genetic diversity, infection dynamics, and viral fitness, we highlight recombination as a key, but nuanced, driver of viral evolution. These findings not only deepen our understanding of viral recombination but also carry important implications for future genomic surveillance, such as to identify periods of heightened risk posed by recombinant variants and to guide public health interventions.

## IV. Materials and methods

### Datasets

We evaluated the genetic diversity, viral fitness, and recombination events using an UShER-based mutation-annotated tree (MAT)^34^ constructed on December 25^th^ 2023, containing 15,990,113 SARS-CoV-2 sequences from the GISAID^38^, GenBank^39^, and COG-UK^40^ online databases. The MAT contained 3,781 pre-annotated internal nodes labeled as the lineage roots for the Pango lineages designated up to that date. The methodology for construction and lineage annotation of the MAT is described in McBroome et al.^41^.

Daily infection counts were obtained from the Johns Hopkins University Center for Systems Science and Engineering (JHU CSSE) COVID-19 Dataset^15^ and aggregated into monthly infection counts (Fig. 1b).

### Inference of putative recombination events

RIVET^19^ was used to infer the set of unique recombinant lineages from the MAT. The RIVET software package provides an automated and optimized implementation of the RIPPLES algorithm^10^ to detect evidence of recombination in pandemic-scale phylogenies. We ran RIVET with the default parameter settings, “*--branch-length 3,--num-descendant 5,--parsimony-improvement 3”*, which requires that a putative recombinant lineage must have a minimum branch length of at least 3 mutations, a minimum of 5 descendant samples, and a minimum partial placement parsimony score improvement of 3, in order to be considered. Running RIVET on the December 25th, 2023 MAT resulted in 3,894 unique recombinant lineages. In order to account for potential false-positive recombinants that could represent artifacts of contamination or result from bioinformatic errors instead of true recombination, RIVET applies various quality control (QC) checks to each putative recombinant and reports on these potential quality issues. Upon examining the distribution of QC flags assigned to the set of Pango-designated recombinant lineages identified by RIVET, we observed that recombinants labeled with the *‘PASS’* and *‘Too_many_mutations_near_INDELS’* QC flags were the most prevalent in our dataset. Given that many of the discovered Pango-designated recombinants were labeled with the ‘*Too_many_mutations_near_INDELS’* flag, we ignored this flag as it is likely too stringent (flagging 1,295 unique recombinant lineages) and could compromise sensitivity. All other detected recombinant lineages containing additional QC flags were excluded from the study. Post filtration, we were left with a total of 2,266 high-quality recombinants.

### Inferring emergence dates of ancestral recombinant sequences

To evaluate and interpret the detectable rate of recombination under the conditions of standing genetic diversity and the number of infections, we inferred the approximate emergence dates for all ancestral nodes in the MAT, using the time-tree estimation method Chronumental^35^. We ran Chronumental version v0.0.65 on the MAT with the following command: “*chronumental –tree $tree--dates $metadata--reference_node $reference_node--steps 2000--only_use_full_dates- -use_gpu*”. Since Chronumental requires an input phylogenetic tree in Newick format, the MAT was first converted to Newick format using the ‘*extract*’ subcommand available in matUtils^41^. The accompanying metadata, which contains the collection dates for all the tip sequences in the given tree, was also provided as input to Chronumental. We selected ‘*CHN/Wuhan_IME-WH01/2019|MT291826.1|2019-12-30*’ as a reference sample that was close to the root of the MAT for Chronumental to compute dates, since erroneous sample collection dates in the metadata can impact the accuracy of date inferences. Additionally, we provided the “*--only_use_full_dates”* option to ensure only samples with collection dates to the precision of a day would be considered. Chronumental defaults to a minimum serial interval of 3 days, and infers a starting molecular clock rate using the root-to-tip (RTT) regression method. The Chronumental job performed 2000 steps for the stochastic variational inference (SVI) and was run on a graphics processing unit (GPU) equipped server. Using the internal node date estimates output from running Chronumental, we labeled each ancestral recombinant lineage in our dataset with its respective date and binned each unique recombination event into one of 37 monthly intervals between February 2020 to February 2023 (Fig. 1b, 2b). We validated that the Chronumental-inferred recombinant emergence dates correlated well with the date of the earliest descendant recorded in the sequence metadata for each recombinant ancestor (Fig. S1). For all experiments involving recombinant fitness (in Fig. 2, 3, 4, S3, S4), sample dates were used to bin sequences into monthly collections of samples, which were used, with the available fitness data, to determine various fitness quantities, such as average and median circulating fitness and percentiles of circulating sample fitness for a given month.

### Defining standing genetic diversity

We use the phylogenetic entropy index^18^ as a metric to capture the circulating genetic diversity available to recombining sequences at a given time. We decided to use a phylogenetic metric, since it accounts for intra-lineage diversity, unlike some other metrics, like co-infection rate and co-circulating lineages^42^. We calculated the standing genetic diversity for each month during a three-year period from February 2020 to February 2023 (Fig. 1b). The MAT was first partitioned into 37 monthly subtrees containing the samples collected for the given month, using the *extract* subcommand in the matUtils software package^41^. The phylogenetic entropy index, H_p_, which was calculated for each of the monthly subtrees, is defined below:

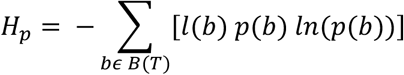

In the above equation, *l*(*b*) is the length of branch *b*, *p*(*b*) is the proportion of the total samples in the subtree that are descendants of branch *b,* and *B*(*T*) is the set of all branches in the given MAT subtree. The quantity H_p_ will increase with the greater diversity of samples in the subtree.

### Examining the fitness of recombinant lineages

To evaluate the fitness implications of a recombination event, we utilized the PyR_017_ model, which provides ranked scoring of thousands of amino acid mutations based on their relative contribution to lineage fitness. We used the amino acid scores from the September 9th, 2023 PyR_0_ model output downloaded from the Broad Institute *pyro-cov* GitHub repository: https://github.com/broadinstitute/pyro-cov. Since PyR_0_ assumes a linear-additive model, we calculated the relative fitness of each recombinant lineage in our dataset by summing the ranked score of each amino acid mutation present in the given lineage. We extracted the set of nucleotide mutations for each recombinant lineage from the MAT and translated each nucleotide mutation to its respective amino acid mutation. Both UShER and PyR_0_ ignore insertions and deletions. Additionally, synonymous mutations, mutations involving codons with ambiguous nucleotides (“N”), and amino acid mutations not scored by the PyR_0_ model were excluded from lineage-level fitness score calculations. Therefore, the relative fitness advantage R/R_a_ represents the relative fitness advantage of the given lineage over the reference SARS-CoV-2 genome. For experiments considering the fitness progression of recombinant lineages over time (Fig. 2), the descendant cluster size for each lineage was extracted from the MAT.

We investigated the relationship between genome divergence of recombining parental sequences and the relative fitness of the child recombinant lineage (Fig. 3b). The divergence of parental genomes was calculated as the Hamming distance between the two sets of nucleotide mutations. The average parental divergence for recombinant lineages in our dataset was approximately 23. When defining the relative fitness of recombinant lineages as their fitness advantage over either of the parental lineages (Fig. 3b), the average fitness of recombinant lineages in our dataset is 0.937, with a standard deviation of 0.138. In comparison, we computed the average fitness of single-nucleotide substitution mutations across all samples in the MAT to be 1.009 with a standard deviation of 0.027. The fitness of each unique recombinant lineage was contextualized into a percentile of all sample circulating fitness for the month that the given recombinant lineage emerged (Fig. 3b,c). Additionally, we considered an alternative definition of relative fitness, in which the fitness of the recombinant lineage is normalized by the minimum fitness of its two parental lineages (Fig. 3c).

To further explore the relationship between parental divergence and recombinant fitness, we selected a handful of recombinant lineages as representative case studies for various outcomes of recombination (Fig. 4). The local phylogenetic tree topology for each recombinant lineage and its parental lineages was examined using the Taxonium^43^ desktop client application version v2.0.223.

### Evaluation of concordance between PyR_0_ and CoVFit fitness models

Since the PyR_0_ model does not account for the potential of epistasis when inferring viral lineage fitness, we considered CoVFit^37^ as an alternative model for determining viral fitness of recombinant lineages. CoVFit is a protein language model that infers fitness as the effective reproduction number (R_e_), by only considering the Spike protein region of SARS-CoV-2 sequences. We chose to evaluate viral fitness using the CoVFit model due to its design to incorporate epistasis and the ease-of-use of the CoVFit CLI. We downloaded the CoVFit CLI executable version “*covfit_cli_20241007*” from the CoVFit GitHub repository (https://github.com/TheSatoLab/CoVFit) and ran the following command to run inference: *“./covfit_cli--input all_spike_translated.fasta-outdir output/ --fold 3 --dms --batch 16 --gpu”.* The input FASTA file, “*all_spike_translated.fasta”*, contained all 6,134 extracted and translated spike protein sequences from unique recombinant lineage trios (recombinant, donor, acceptor) in our dataset. We used the default CoVFit parameters, including a sequence batch size of 16, model instance 3, and utilized a GPU to perform inference. We compared the PyR_0_ fitness scores against the CoVFit fitness (R_e_) scores for each of the recombinant lineages in our dataset (Fig. S2). We computed the Pearson correlation coefficient between the two fitness metrics and fit a regression model to the dataset to compute the R-squared value (Fig. S2).

### Phylogenetic Tree Visualization

We used the Taxonium^43^ desktop application version v2.0.223 to view the phylogenetic tree for several recombination/donor/acceptor trios for the December 25th 2023 MAT. This Taxonium tree was generated from the MAT and associated sample metadata using the *taxoniumtools* Python package and CLI to convert the MAT Protocol Buffers binary format to the Taxonium jsonl format. The “usher_to_taxonium” command was run with the “--name_internal_nodes” option to include MAT internal node labels that match the RIVET recombinant inference results. The full command and script to generate the Taxonium tree is provided in the accompanied GitHub repository. We downloaded SVG images of the local phylogenetic tree for each recombinant trio from Taxonium (Fig. 4). The branch lengths of these phylogenetic tree images are to approximate scale, due to being slightly resized using Adobe Illustrator.

## Data and Code Availability

All code developed in this study is freely available under the MIT license at https://github.com/TurakhiaLab/SARS2_RecombinationAnalysis, which provides scripts and Jupyter notebooks to reproduce all experiments and figures from this study. For purposes of reproducibility and ongoing recombinant surveillance and study, several metrics and useful functionality from this study have been contributed upstream to RIVET since version v0.3.0 and is freely available under the MIT license at https://github.com/TurakhiaLab/rivet.

## Data Availability

All data used in this study was retrieved from online databases. The SARS-CoV-2 mutation-annotated tree (MAT) used for analysis was constructed on December 25, 2023, using genomic sequences obtained from the GISAID, GenBank, and COG-UK databases, and is made available through the UShER database. Daily infection counts were sourced from the Johns Hopkins University Center for Systems Science and Engineering (JHU CSSE) COVID-19 Dataset (https://coronavirus.jhu.edu/). The PyR0 model outputs used for fitness evaluation were downloaded from the Broad Institute pyro-cov GitHub repository (https://github.com/broadinstitute/pyro-cov). CoVFit fitness scores were generated using the CoVFit CLI executable available at https://github.com/TheSatoLab/CoVFit. All code developed for this study is freely available under the MIT license at https://github.com/TurakhiaLab/SARS2_RecombinationAnalysis, which includes scripts and Jupyter notebooks to reproduce all experiments and figures.

## Acknowledgements

We thank the authors and developers of the PyR_0_ model, especially Jacob Lemieux and Benjamin Kotzen, for providing updated mutation fitness results and for their assistance with PyR_0_. We thank Josh Levy and Kristian Andersen for helpful discussions. We also thank all data contributors, authors, and their originating laboratories responsible for obtaining the specimens and their submitting laboratories for generating the genetic sequence and metadata, and sharing via the GISAID Initiative, INSDC, COG-UK, and the China National Center for Bioinformation, on which this research is based. This work was supported by funding from the Hellman Fellowship (to Y.T.) and the Amazon Research Award (Fall 2022 CFP).

## Competing interests

J.O.W. and Y.T. have received contracts from the Centers for Disease Control and Prevention (CDC) for viral molecular surveillance, not directly related to this work. J.O.W. has also provided compensated expert testimony on SARS-CoV-2 and the COVID-19 pandemic.

