## Supplementary Figure for "Genetic Diversity Drives the Rate and Fitness Jumps of Detectable SARS-CoV-2 Recombination"

### Supplementary Figures:

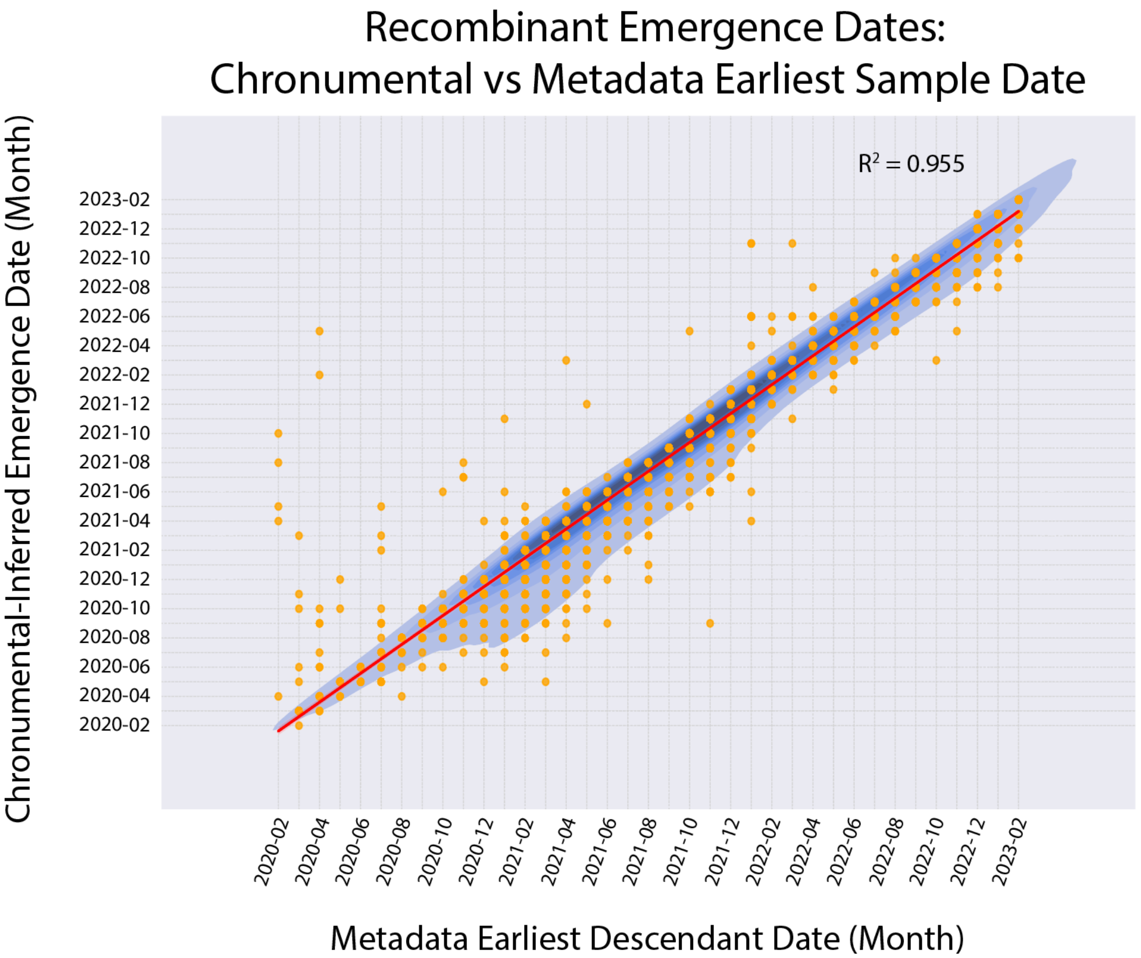


**Figure S1: Inferring Recombinant Emergence Dates with Chronumental versus Earliest Descendant Sample Metadata Date**

A comparison of RIVET-inferred recombinant emergence dates estimated using Chronumental against the earliest recorded date (in the sequence metadata) from the set of descendants for each recombinant.

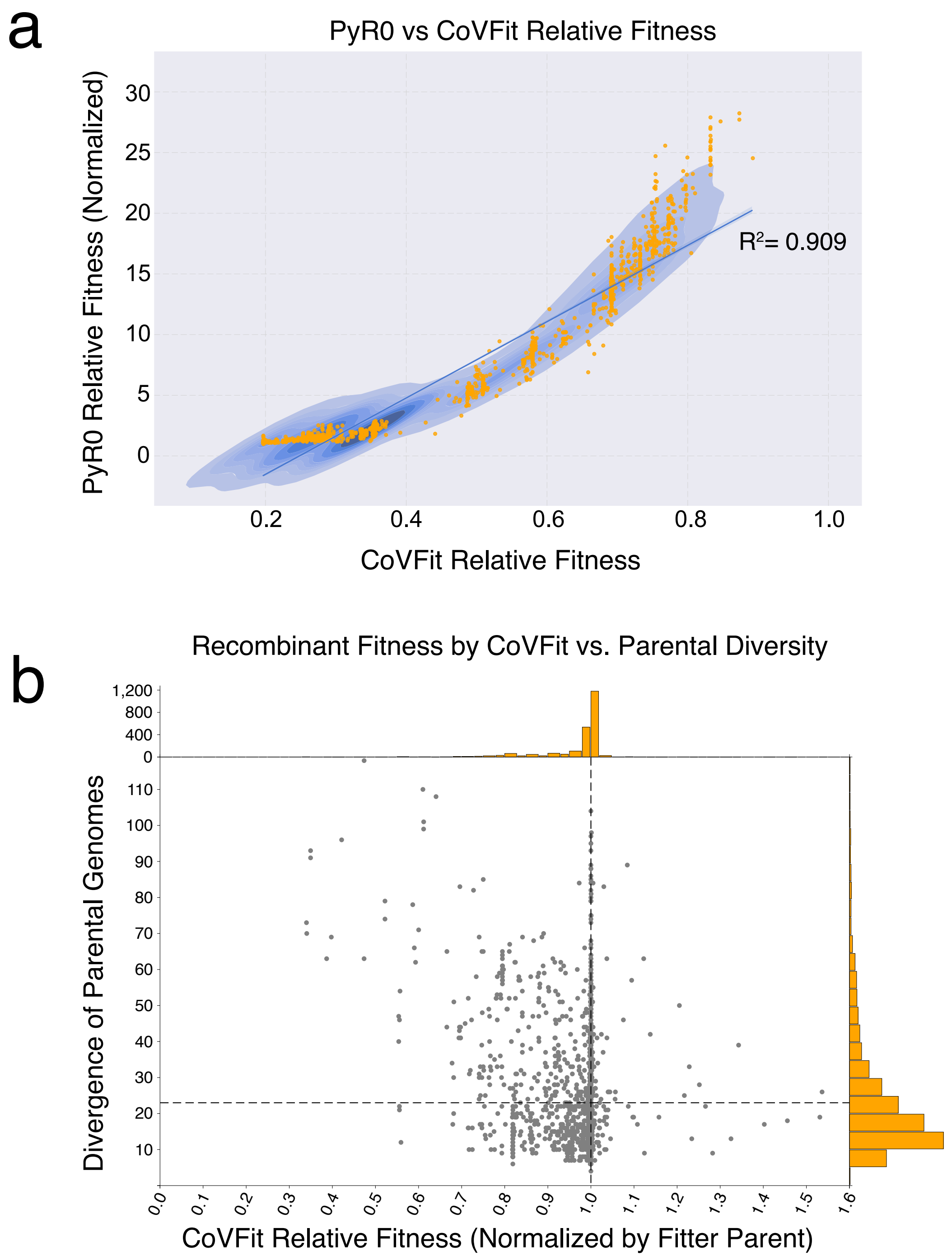


**Figure S2: Comparing recombinant relative fitness in PyR$_0$ vs CoVFit models**

(**a**) We compare the relative fitness scores (normalized by the fitter parent) assigned by the PyR$_0$ and CoVFit models for all the recombinants in our dataset. (**b**) The relationship between relative fitness (using the CoVFit model and normalized by the fitter parent) and the divergence of the recombining parental sequences. The horizontal dashed line denotes the average parental sequence mutational distance.

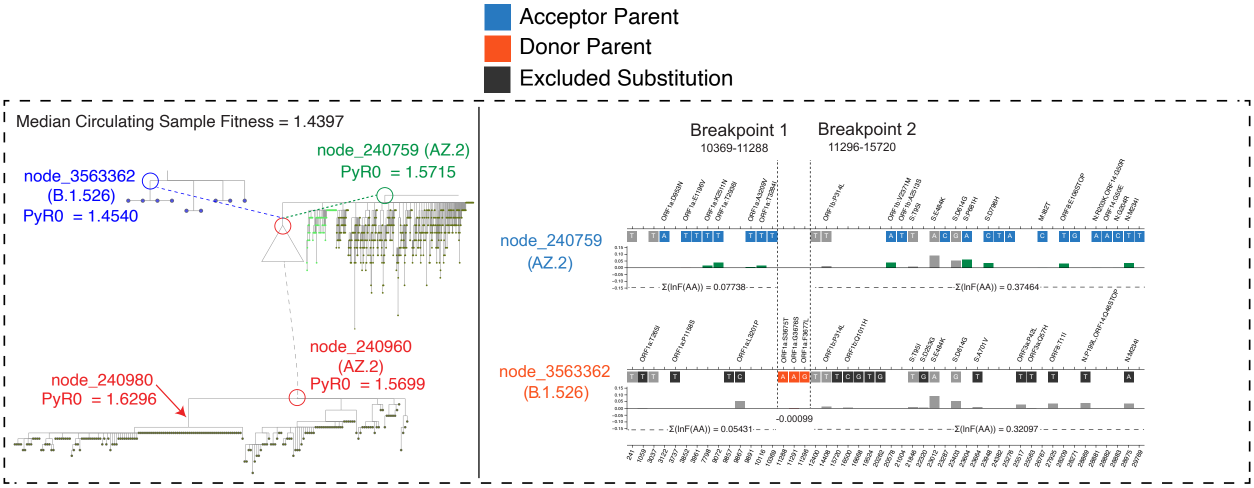


**Figure S3: Case study of a fitness-neutral recombination event from highly divergent parental sequences.** The left panel shows the approximate phylogenetic tree topology of a recombination event inferred at node_240960. The recombination between highly divergent parental lineages, a B.1.526 donor sequence and a AZ.2 acceptor sequence with higher fitness than the median circulating fitness ($PyR_0$), resulted in a near-neutral recombinant with respect to fitter AZ.2 parent. On the right-side panel, the acceptor (blue) and donor (orange) protein-coding, non-synonymous single-nucleotide substitutions are shown. The vertical dashed lines denote the inferred breakpoint intervals for the detected trio. The relative fitness ($PyR_0$) of each amino acid mutation is shown on the track below each nucleotide change, with fitness-increasing (green) and fitness-decreasing (red) mutations highlighted.

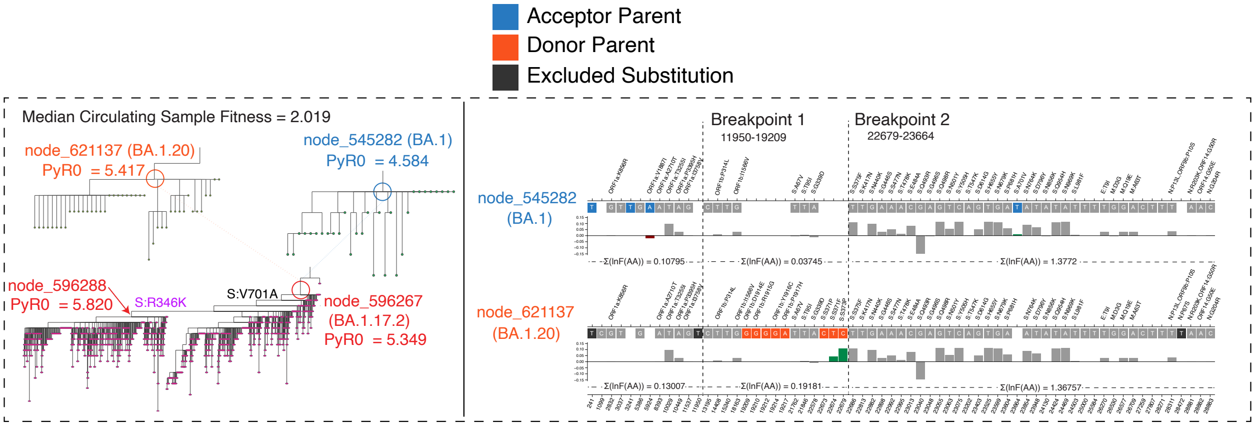


**Figure S4. Case study of a fitness-neutral recombination event from genetically similar parental sequences.** The left panel shows the approximate phylogenetic tree topology of a recombination event inferred at node_596267, between parental sequences from the BA.1 and BA.1.20 lineages. The nearly neutral recombination event (with respect to the fitter parent), introduces new diversity within the spike protein region. Further spike protein substitutions are observed in short succession on the branches immediately following the recombinant node, including a R346K mutation, which $PyR_0$ ranks as the third-fittest mutation. On the right-side panel, the acceptor parent (blue) and donor parent (orange) protein-coding, non-synonymous single-nucleotide substitutions are shown. The vertical dashed lines denote the inferred breakpoint intervals for the detected trio. The relative fitness of each $PyR_0$ amino acid mutation is shown on the track below each nucleotide change, with fitness-increasing (green) and fitness-decreasing (red) mutations highlighted.
